# Personalized structural biology reveals the molecular mechanisms underlying heterogeneous epileptic phenotypes caused by *de novo* KCNC2 variants

**DOI:** 10.1101/2022.02.01.21268115

**Authors:** Souhrid Mukherjee, Thomas A. Cassini, Ningning Hu, Tao Yang, Bian Li, Wangzhen Shen, Christopher W. Moth, David C. Rinker, Jonathan H. Sheehan, Joy D. Cogan, Undiagnosed Diseases Network, John H. Newman, Rizwan Hamid, Robert L. Macdonald, Dan M. Roden, Jens Meiler, Georg Kuenze, John A. Phillips, John A. Capra

## Abstract

**Background:** Next-generation whole exome sequencing (WES) is ubiquitous as an early step in the diagnosis of rare diseases and the interpretation of variants of unknown significance (VUS). Developmental and epileptic encephalopathies (DEE) are a group of rare devastating epilepsies, many of which have unknown causes. Increasing WES in the clinic has identified several rare monogenic DEEs caused by ion channel variants. However, WES often fails to provide actionable insight, due to the challenges of proposing functional hypotheses for candidate variants. Here, we describe a “personalized structural biology” (PSB) approach that addresses this challenge by leveraging recent innovations in the determination and analysis of protein 3D structures.

**Results:** We illustrate the power of the PSB approach in an individual from the Undiagnosed Diseases Network (UDN) with DEE symptoms who has a novel *de novo* VUS in *KCNC2* (p.V469L), the gene that encodes the Kv3.2 voltage-gated potassium channel. A nearby *KCNC2* variant (p.V471L) was recently suggested to cause DEE-like phenotypes. We find that both variants are located in the conserved hinge region of the S6 helix and likely to affect protein function. However, despite their proximity, computational structural modeling suggests that the V469L variant is likely to sterically block the channel pore, while the V471L variant is likely to stabilize the open state. Biochemical and electrophysiological analyses demonstrate heterogeneous loss-of-function and gain-of-function effects, respectively, as well as differential inhibition in response to 4-aminopyridine (4-AP) treatment. Using computational structural modeling and molecular dynamics simulations, we illustrate that the pore of the V469L variant is more constricted increasing the energetic barrier for K^+^ permeation, whereas the V471L variant stabilizes the open conformation

**Conclusions:** Our results implicate *KCNC2* as a causative gene for DEE and guided the interpretation of a UDN case. They further delineate the molecular basis for the heterogeneous clinical phenotypes resulting from two proximal pathogenic variants. This demonstrates how the PSB approach can provide an analytical framework for individualized hypothesis-driven interpretation of protein-coding VUS suspected to contribute to disease.

## INTRODUCTION

The advent of cheaper and more accurate DNA sequencing technologies has enabled the integration of genetic information into diverse areas of medicine. For example, more than 70% of rare diseases are thought to have a genetic cause, and recent efforts have identified the causal variants for thousands of Mendelian diseases ^1–3^. However, causal variants have not been identified for approximately half (∼3000) of known rare genetic diseases ^4–6^, and sequencing often fails to lead to actionable insights, even after expert clinical evaluation through programs like the NIH’s Undiagnosed Diseases Network (UDN) ^7–9^.

Many computational methods have been developed for interpreting variants observed in clinical sequencing ^10–13^. However, they have substantial weaknesses and often disagree ^14–18^. In particular, commonly used tools provide only ill-defined, categorical variant classifications like “pathogenic” and “benign” and fail to propose specific hypotheses about the underlying molecular effects of variants. A prediction that a variant is “pathogenic” is not of much clinical use without a testable prediction of the mechanisms of its pathogenicity, pleiotropic effects and possible insights to treatment.

Motivated by the challenges of variant interpretation, recent advances in experimental approaches for protein structure determination ^19–24^ and recent improvements to the accuracy of prediction, modeling and analysis of native 3D protein structural models ^25–28^, we propose a new variant interpretation paradigm. Our “personalized structural biology” approach focuses on making mechanistic predictions about the effects of the variant(s) observed in patients in the context of their genetic background via computational and experimental evaluation of protein structure and function. We demonstrate the power of this approach on two candidate VUS in *KCNC2*, the gene encoding the homo-tetrameric voltage gated potassium channel Kv3.2, one variant from an individual with an unsolved epilepsy-like disease enrolled in the UDN and another variant from a recent case report ^29^ with epilepsy-like phenotypes; however no functional validation was done for the variant.

Developmental epileptic encephalopathies (DEE) are a group of devastating disorders in which epileptic activity contributes to cognitive and behavior impairment in addition to underlying developmental pathologies ^30,31^. Genetic etiologies are thought to be the cause of a substantial proportion of these DEE cases, and with recent advances in genetic testing technology, many DEE variants have been discovered. The underlying genetic mechanisms are diverse ^32^, but defects in neuronal ion channels are thought to be a common cause of DEE. For example, the initial discovery that Dravet syndrome is caused by variants in *SCN1A* ^33^ has been followed by the demonstration that variants in many voltage-gated potassium (Kv) channels can cause DEE^34^. The largest family of these channels is the Kv family, with 12 subfamilies whose alpha subunits are encoded by approximately 40 genes. The Kv3 subfamily influences rapid firing of inhibitory interneurons in the central nervous system ^35^. The general mechanism of Kv3.2 channel gating is thought to be similar to other closely related voltage-gated potassium ion channels. Kv channels consist of four homologous subunits, with each monomer having six transmembrane helical domains (S1-S6). S1-S4 form the voltage sensing domain (VSD) and S5-S6 from all four subunits form the membrane pore. A linker domain between S4 and S5 connects the VSD to the pore forming units ^36–40^. The VSD undergoes conformational changes between the open and closed state of the channel ^41^, and the coupling between the S4-S5 linker and the S6 pore forming unit is responsible for the voltage dependent gating of the channel ^42,43^. With a pronounced inward movement of the positively charged S4 voltage sensor, the S4-S5 linker is pushed downwards, which causes the S6 helix to constrict the pore, thus closing the channel ^39^. This gating mechanism is made possible by the presence of a Proline-Valine-Proline (PVP) motif, which acts as a hinge domain in the S6 helix. The hinge domain allows the S6 pore-forming helix to kink at the flexible PVP motif to open and close the channel pore.

Other members of this subfamily have been implicated as a potential causes of DEE and other epilepsy-like symptoms with discoveries of variants in genes encoding Kv3.1 and Kv3.3 ^44– 46^. More recently variants in *KCNC2*, which is highly expressed in GABAergic interneurons in the CNS, have been suggested to be linked to DEE-like phenotypes, with possibly dominant negative effects ^29,47,48^. However, the links and their mechanisms are yet to be established.

Our work makes four main contributions. First, we demonstrate via expression and electrophysiology analyses that the two candidate *KCNC2* variants (p.V469L, p.V471L) have loss-of-function and gain-of-function effects, respectively, despite both affecting the essential hinge region of Kv3.2 responsible for channel gating. Second, our protein structural modeling and molecular dynamics simulations rationalize the mechanistic basis for the phenotypic heterogeneity of these variants. Third, our results combine to validate links between *KCNC2* variants and heterogenous DEE phenotypes. Finally, our analyses provide a blueprint for integrating genetics, expression analysis, electrophysiology, and protein structural modeling to develop mechanistic understanding of the molecular effects of *de novo* variants in rare disease.

## RESULTS

### Undiagnosed Diseases Network patient with DEE-like symptoms

A child (3-8 years old) at the Vanderbilt University UDN site presented with DEE-like phenotypes, including multiple types of refractory seizure and global developmental delay. At a very young age, they developed generalized tonic clonic seizures, and was diagnosed with Lennox-Gastaut syndrome, a severe form of DEE. However, they continued to have frequent myoclonic absence seizures and occasional generalized tonic clonic seizure. The IRB of the National Institutes of Health (NIH) gave ethical approval for this work (protocol 15HG0130). The participants’ or their legal representatives’ have consented to IRB 15HG0130 and allowed the results of this research work to be published. A full clinical report is available upon contacting the corresponding authors.

Initial sequencing of the individual on an epilepsy gene panel through Athena covered *ARHGEF9, ARX, CDKL5, CNTNAP2, FOXG1, GABRG2, GRIN2A, KCNT1, MECP2, NRXN1, PCDH19, PNKP, RNASEH2A, RNASEH2B, RNASEH2C, SAMHD1, SCN1A, SCN1B, SCN2A, SCN8A, SCN9A, SLC25A22, SLC2A1, SLC9AC, SPTAN1, STXBP1, SYNGAP1, TCF4, TREX1, UBE3A, ZEB2*. The only potentially significant finding was a heterozygous c.2985G>C variant in *GRIN2A*. Deletion analysis of *SCN1A* was negative as well. Secondary findings, metabolic screens, and mitochondrial DNA sequencing were also negative.

Following the negative epilepsy panel result, WES revealed a candidate variant in a voltage-gated potassium channel Kv3.2, *KCNC2* c.1405G>T (p.V469L). Sanger sequencing confirmed this variant. This variant was not seen in either of his parents, and therefore it was presumed to be *de novo*. This variant is in the conserved hinge motif of the channel which is critical for channel gating (**Figure 1**). The potential relevance of this variant is supported by another recently reported discovered candidate heterozygous variant also located in the hinge domain of *KCNC2* (c.1411G>C, p.V471L) only two amino acids away from V469L variant found in our UDN subject ^29^. Our UDN subject and the previously reported case shared the phenotypes of DEE, seizures refractory to medications, developmental delay, and microcephaly. However, their phenotypes differed in that the reported case also had complete absence of speech, dystonia, decreased myelination around frontal and occipital horns of the lateral ventricles, spastic tetraplegia, myoclonic jerks, and opisthotonos attacks.

**Figure 1:**
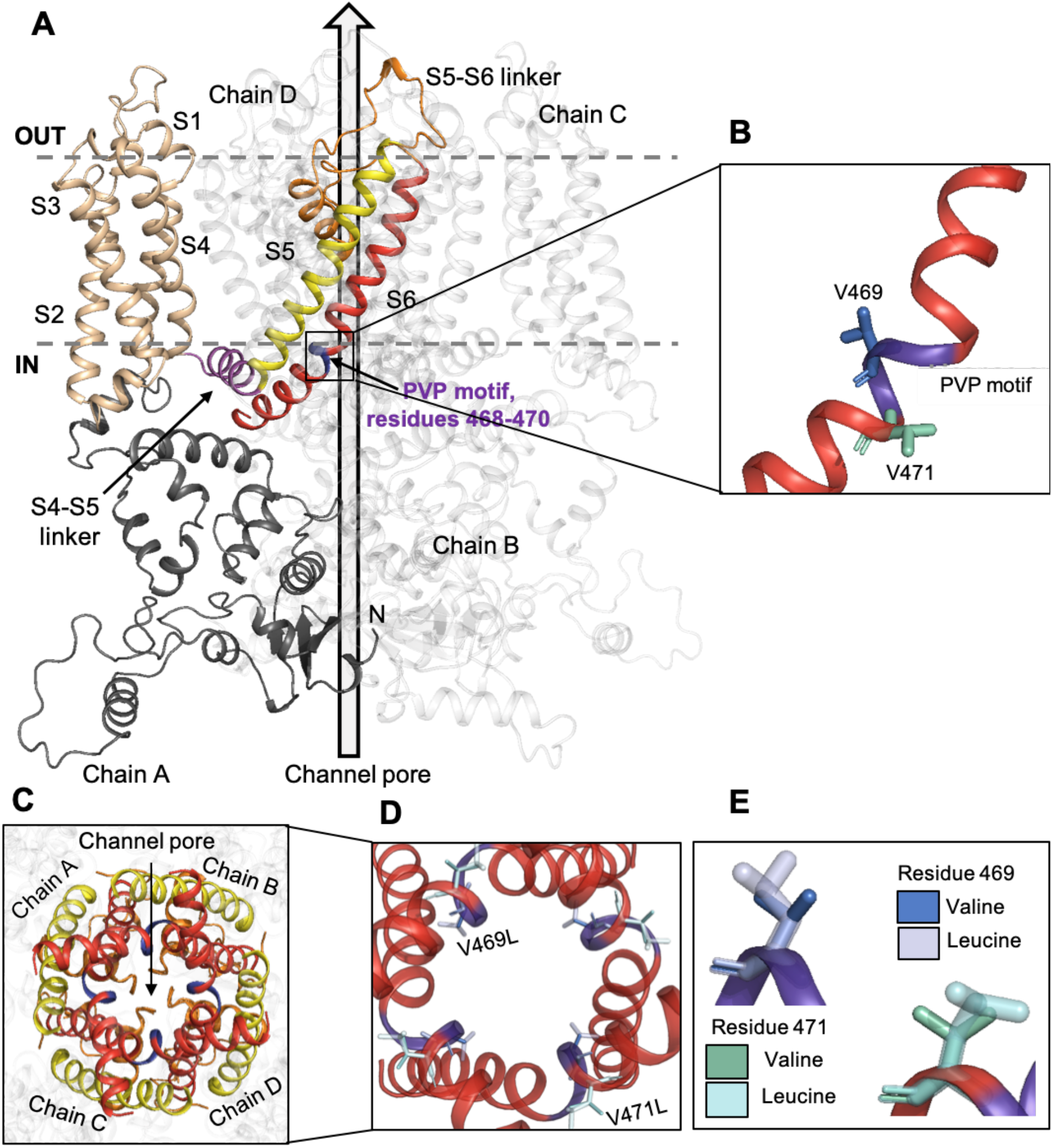
Candidate pathogenic variants in *KCNC2* are nearby, but have different structural contexts in Kv3.2. **(A)** Homo-tetrameric structure of Kv3.2. The complete structural model of Kv3.2 (*KCNC2*) was generated using RosettaCM from Kv1.2-Kv2.1 paddle chimera channel (PDB ID: 2R9R, 42.8% sequence identity). Four homologous subunits form the tetrameric channel pore structure; chain A is shown in color. Each monomeric subunit has intracellular N terminal domain (black) and six transmembrane helical domains. S1-S4 form the voltage sensing domain (VSD, beige). The S4-S5 linker (magenta) is the force transducer between the VSD and the channel pore, formed by the S5 (yellow) and the S6 (red) helices. The S5-S6 linker (orange) acts as the selectivity filter, allowing only potassium ions through the channel. The patient variant (V469L) is located in the PVP motif (purple; residues 468-470) which acts a hinge domain facilitating channel gating. The previously discovered variant (V471L) suspected to also cause DEE-like symptoms is located adjacent to the PVP motif. **(B)** A view of the carbon backbone of the S6 helix, showing the PVP hinge region. The valines at positions 469 (blue) and 471 (green) are shown. **(C)** A view of the channel pore formed by the tetrameric structure of Kv3.2, showing the selectivity filter (orange) and the hinge domain (purple). **(D)** A closer view of the channel pore with reference amino acids valine at positions 469 (deep blue) and 471 (teal) shown, alongside variant leucine residues (light blue and cyan). Residue 469 extends into the pore, while residue 471 faces away from the pore. **(E)** A view of the carbon backbone of the native (valine) and substituted (leucine) amino acids at positions 469 and 471.

To evaluate the evidence for these VUS and propose specific functional hypotheses, we assessed the effects of these variants on protein expression, structure, and function with experimental and computational methods.

### Structural modeling suggests distinct functional effects for candidate KCNC2 variants

To evaluate the potential effects of the *KCNC2* variants at the molecular level, we constructed a homology model of its tetrameric structure based on a high-resolution structure of the Kv1.2-Kv2.1 paddle chimera channel (PDB ID: 2R9R) ^42^ using the Rosetta molecular modeling suite (Methods) ^49^.

The homo-tetrameric Kv3.2 model, with six transmembrane helical domains (S1-S6) is shown in **Figure 1A**. The PVP motif ranges between residues 468 and 470 on the S6 helix and facilitates channel gating. The variants of interest p.V469L and p.V471L are adjacent to the PVP motif (**Figure 1B**). The channel pore is formed by the S5 and S6 helices of all four chains together (**Figure 1C**), and the PVP motifs on all four helices act together for channel gating. This region is almost entirely conserved among vertebrates (**Figure S1A**). Furthermore, previous studies have shown that altering the central hydrophobic residues in Kv1.1 channels from valine to isoleucine affects channel kinetics, stability, and conformational dynamics ^50^.

The UDN subject’s variant (p.V469L) results in a conservative substitution, which changes the hydrophobic valine at the core of the PVP motif to leucine, another hydrophobic amino acid. However, the p.V469L variant is predicted to have a deleterious effect on the protein by commonly used variant effect prediction tools like CADD and GERP (**Figure S1B)**. The residue at position 469 faces into the channel pore (**Figure 1D)**, and while conservative amino acid substitutions do not usually have severe effects, in this case, the bulkier leucine amino acid (**Figure 1E**) at the center of the PVP motif on the hinge domain could influence ion transfer. We hypothesize that it could sterically obstruct the pore resulting in a decreased pore radius and slower kinetics of channel gating. The steric hindrance of the curving of the S6 helix could lead to fewer ions passing though the pore.

The second recently reported candidate *KCNC2* variant ^29^ (p.V471L) is immediately adjacent to the PVP motif (**Figure 1A**); however, the structural context of this variant in our model suggests potentially different effects from that of our UDN subject’s p.V469L variant. Residue p.V471 faces away from the pore (**Figure 1D**), and therefore, the substitution of the bulkier amino acid leucine (**Figure 1E**) is less likely to lead to a decrease in the pore radius as the residue faces outward. In this case, we hypothesize that the molecular effect of the p.V471L variant would widen the pore and increase its tendency to remain open, leading to more ions passing through than normal and thus a gain-of-function phenotype. We also predict that the channel gating will be affected by the bulkier leucine residue; however, not to the extent of the p.V469L variant since p.V471L faces away from the channel pore.

### V469L causes loss of channel function, while V471L causes gain of function

To quantify the effects of the candidate variants we quantified the electrophysiological function of Kv3.2, potassium channel currents for WT Kv3.2 and the two disease causing variants (p.V469L and p.V471L) in CHO cells. The proteins were expressed in a homo-tetrameric model, with all four chains carrying the variant, in each case. The WT form of Kv3.2 showed a very fast deactivation, in accordance with previously characterized behavior of the Kv3.2 channel (**Figure 2A, S2**). *KCNC2* is primarily expressed in the brain, where its product Kv3.2 contributes to the fast repolarization of action potentials in neurons of the central nervous system ^35,51,52^. Therefore, short spike duration and rapid deactivation of Kv3.2 channels are important for normal physiology.

**Figure 2:**
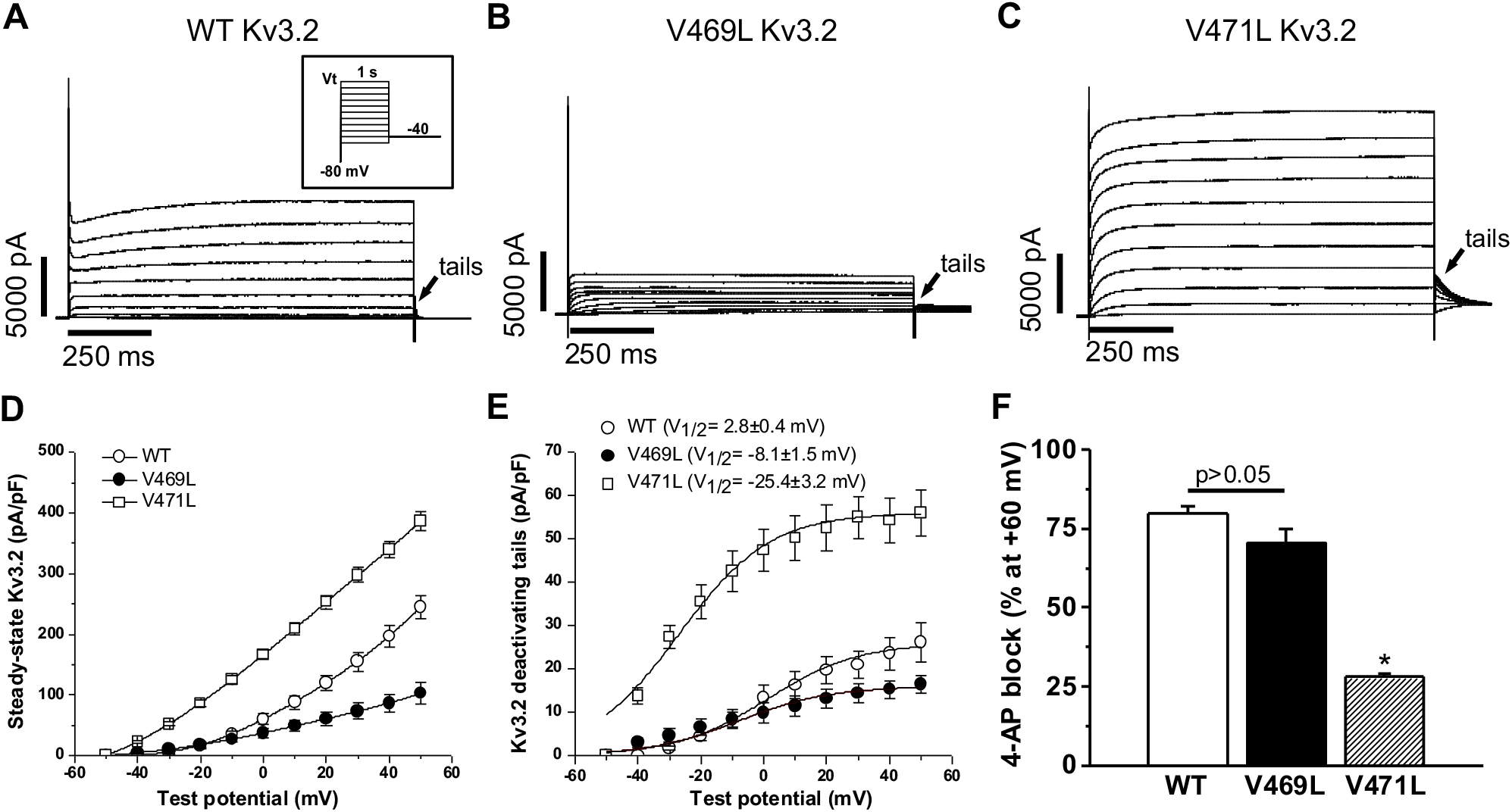
Candidate Kv3.2 variants cause loss and gain of channel function. **(A)** Representative potassium current traces for wild type (WT) Kv3.2 in CHO cells recorded by voltage clamp. WT shows the characteristic current amplitude and fast deactivation (short deactivating tail currents). **(B)** Current traces for the V469L variant show lower peak current and very slow deactivation. **(C)** Current traces for the V471L variant demonstrate much higher current and moderately slowed deactivation compared to WT. Each voltage clamp used the protocol shown in the insert of panel A. The deactivation tails are compared in greater detail in **Figure S2A-C. (D)** Current vs. steady-state voltage (I-V) curves for WT Kv3.2, V469L, and V471L. The V469L variant showed a much lower current at steady-state, while the V471L variant showed increased current at steady-state. **(E)** Current vs. voltage plots for the tails for each variant. V469L has a slight negative shift (∼10 mV), while V471L has a large negative voltage shift (∼28 mV). Each group considered 6 to 10 cells. **(F)** Percentage of channel activity (steady-state current) blocked by 200 µ M 4-aminopyridine (4-AP), a known voltage gated potassium channel blocker, for WT, V469L, and V471L. The WT and V469L Kv3.2 were similarly blocked (p > 0.05, n=6 for each), but V471L was resistant to 4-AP blockage (p < 0.0001, n=6). **Figure S3A-C** shows the protocol and representative traces for each variant. Altogether, these data demonstrate loss- and gain-of-function effects on channel activity for V469L and V471L, respectively.

The electrophysiological profiles for the p.V469L variant (**Figure 2B, S2)** compared to the WT Kv3.2 had a lower peak current and much longer deactivation tails. The peak current for the p.V469L mutant was less than half that of WT (**Figure 2D**), with a much slower deactivation and a slight negative shift of ∼10mV (**Figure 2E)**. In contrast, the electrophysiological profile for p.V471L (**Figure 2C, S2**) showed a much higher peak current than WT, with a longer deactivation tail. The p,V471L peak current was 1.5 times that of the WT channel (**Figure 2D**), with a moderately slower deactivation, but a drastic negative shift of ∼28mV (**Figure 2E**). The very slow deactivation and slightly negative voltage shift observed for p.V469L and moderately slow deactivation and dramatically negative voltage shift for p.V471L align with the structural hypothesis of loss-of-function and gain-of-function phenotypes, respectively. For each variant, the same amount of plasmid was injected, and the behavior of the proteins at their native levels of expression was analyzed.

We further characterized the effect of the variants on channel function by administration of the voltage-gated potassium channel blocker 4-aminopyridine (4-AP). Kv3.2 is very sensitive to 4-AP ^53–55^. 4-AP is known to approach the channel lumen from the cytoplasmic side ^53,56,57^; and bind the open channel weakly. Once bound, the channel becomes biased towards the closed state ^56^, and 4-AP binds strongly to the closed conformation, blocking the channel.

Interestingly, 4-AP blocked the channel activity similarly for the WT and p.V469L Kv3.2 (**Figure 2F, S3**). Both experienced >70% decreases in activity (p > 0.05, n=6). In contrast, the gain-of-function p.V471L variant was resistant to 4-AP compared to WT and p.V469L (p < 0.0001 for both, n=6), showing a less than 30% reduction in activity (**Figure 2F, S3**). This could be due to the V471L variant stabilizing the channel in the open conformation, thereby making 4-AP less effective in closing the channel and less likely to bind. These results further support the contrasting loss- and gain-of-function mechanisms we propose for p.V469L and p.V471L.

### V469L expression is lower while V471L expression is higher than wild type

Rare pathogenic protein-coding variants, in addition to causing changes to protein structure and molecular function, can also lead to altered protein expression in cells. We compared the levels of protein present in cells for the WT and two Kv3.2 variants with an Immunoblot analysis (Western Blot).

The homomeric V469L Kv3.2 was present at less than half the amount for homomeric WT, while the levels for homomeric V471L Kv3.2 were greater than one and a half times that of WT (**Figure 3**). Thus, expression differences likely contribute to the loss- and gain-of-function effects for the two variants, respectively. However, while the expression levels could cause the observed differences in the peak currents for the two variants (**Figure 2**), differences in protein levels alone cannot explain the slowed deactivation dynamics of the V469L channel. Thus, both differences in the molecular function and expression levels of these *KCNC2* variants contribute to their phenotypic effects.

**FIGURE 3:**
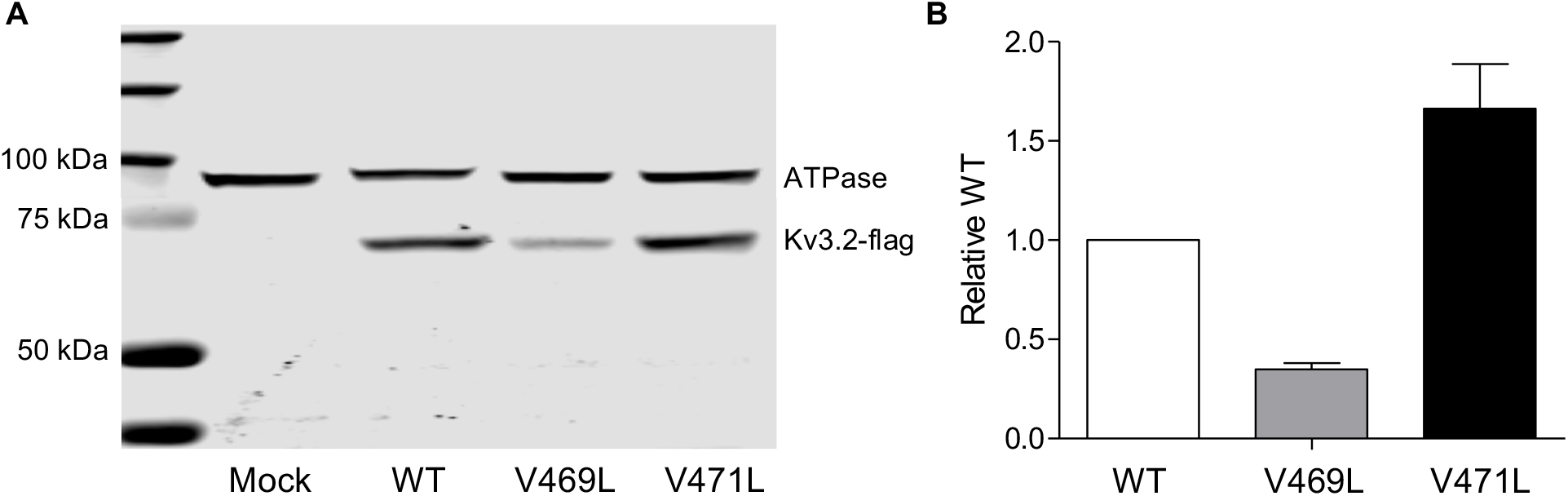
Candidate *KCNC2* variants modify Kv3.2 expression levels. **(A)** Western blot showing expression of WT Kv3.2, V469L and V471L variants in CHO stable cells. The mock well shows only the ATPase antibody tag, while the WT, V469L and V471L labeled wells have the corresponding version of Kv3.2 loaded. Kv3.2 has a molecular weight of ∼70 kDa and shows up as one band, below the 75 kD marker, thus confirming the presence of the protein in its native state. The V471L band is larger and more intense than WT, while V469L is faint. This suggests higher protein levels for V471L and lower levels for V469L compared to WT. **(B)** Protein expression estimated from Western blot band intensity. The protein levels for V469L were roughly half that of the WT, and the proteins levels for the V471L were more than one and a half times that of WT. These results support a loss-of-function phenotype for V469L and a potential gain-of-function phenotype for V471L.

### V469L constricts the channel pore in molecular dynamics simulations increasing the energetic barrier for K^+^ ion permeation

To explore the molecular basis for the functional changes caused by V469L and V471L, we performed molecular dynamics simulations of these ion channel variants and of WT Kv3.2 in POPC membranes. Each system was simulated for more than 1μs in total. **Figure 4A-C** displays simulation snapshots of the three ion channel systems and **Videos V1-V6** show representative MD trajectories. We observed that the inner pore helices in V469L moved closer together at their hinge motif sites such that the pore became more constricted and fewer water molecules were able to enter the inner channel cavity through the cytosolic gate. This effect was most likely driven by increased attractive interactions between leucine 469 residues on adjacent and opposite S6 helices that led to a ‘de-wetting’ of the channel pore. Calculation of the pore radius along the channel axis (z-axis) (**Figure 4D**) confirmed that the K^+^ ion permeation pathway in the V469L channel is more constricted compared to WT and V471L Kv3.2 channels.

**FIGURE 4:**
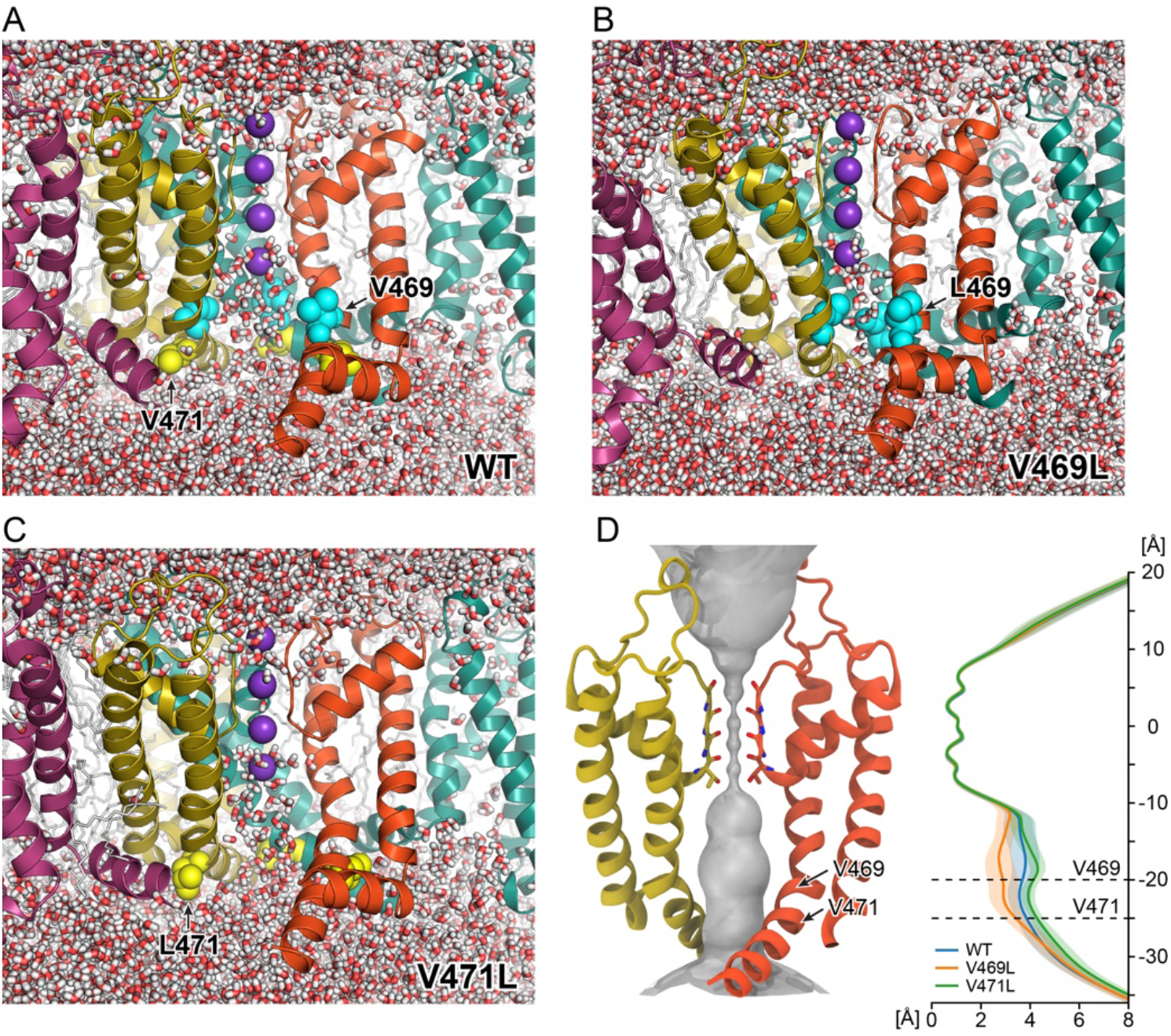
The channel pore of Kv3.2 V469L becomes constricted in MD simulations whereas the pore of V471L adopts a stable open conformation. **(A)** – **(C)** Snapshots from MD simulations of Kv3.2 WT, V469L, and V471L. The entire MD trajectories are shown in **Videos V1-V6**. For each protein, the channel-membrane system was simulated in four replicas with a total simulation time of more than 1μs. The protein is represented as ribbon with each chain shown in a different color. One domain in the front is not shown to better see the channel cavity. The amino acids at positions 469 and 471 are depicted as spheres and colored cyan and yellow, respectively. Water molecules are shown as sticks (red-white). **(D)** Surface representation of the pore radius of Kv3.2 WT (left) and 1D pore radius profiles (right) along the channel axis (z-axis) for WT, V469L, and V471L. The solid line and shaded area represent the average radius and standard deviation from four independent MD replicas.

Furthermore, we noticed small but distinct differences of the backbone structure at residue 469 and preceding residues (**Figure S4, Table T1**). By contrast, the pore radius of the V471L channel was slightly wider than that of WT Kv3.2 indicating that V471L adopted a stable open conformation in MD. One possible explanation for this observation is the difference in the types of interactions made by residues at positions 469 and 471. While L469 side chains are oriented towards each other and towards the pore, L471 residues are oriented outward and interact with residues on S5 in the same subunit and with residues on S6 in a neighboring subunit. The largest number of atom contacts of L471 are made with Y480 on an adjacent S6 helix (**Figure S5**). In the variant, the number of heteroatom contacts (within 4Å radius) for the L471-Y480 interaction more than doubled relative to the V471-Y480 interaction in WT Kv3.2 (average of ∼3.2 contacts in WT Kv3.2 to ∼7.0 contacts in the V471L channel). This finding offers a plausible explanation for how this amino acid change at position 471 leads to stabilization of the open channel conformation.

To assess the energetic cost more directly for K^+^ ion permeation in WT Kv3.2 and both channel variants, we used umbrella MD simulations and calculated the potential of mean force (PMF) for moving a K^+^ ion from the cytosolic site of the channel through the S6 helix gate into the water-filled cavity below the selectivity filter (**Figure 5A**). Compared to WT, the energetic barrier for ion transfer of the V469L variant increased by ∼0.8 kcal/mol (**Figure 5B**). The V471L variant, however, required an energy for ion transfer similar to WT. The highest peak in the PMF and the V469L-specific energy increase occurred at position P470. This indicates that V469L, but not V471L, constricts the channel pore at the PVP hinge region, which is in line with our pore radius measurements.

**FIGURE 5:**
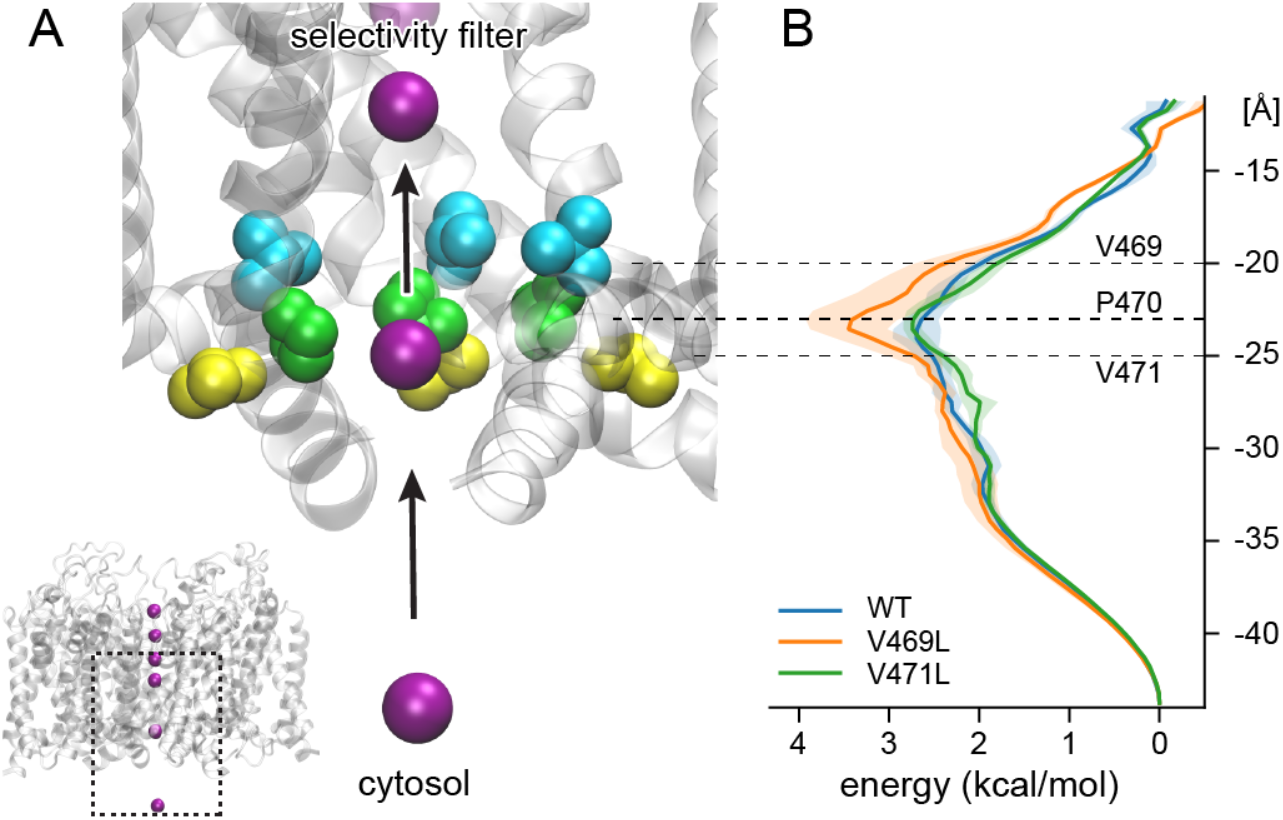
V469L increases the energy required for K^+^ ion transfer through the cytosolic gate of Kv3.2 compared to WT and V471L. **(A)** We estimated the energy required to transfer a K^+^ ion (purple sphere) through the cytosolic channel gate, from the bulk solvent into the cavity below the selectivity filter by umbrella MD simulation. A close-up view of the gate and aqueous cavity of WT Kv3.2 channel is shown. The subunit in the front is not depicted to better see the K^+^ permeation pathway which is lined by residues on S6. The side chains of reference amino acids V469, P470, and V471 are drawn as cyan, green, and yellow spheres, respectively. **(B)** 1D PMF of K^+^ transfer through the channel for Kv3.2 WT, V469L, and V471L. The solid line and shaded area represent the average PMF and standard deviation of four independent MD simulations. The V469L variant induces a greater energetic barrier to ion transfer compared to WT and V471L. The increased energetic requirement is focused on the conserved P470 residue in the hinge region. This supports the relevance of the disruption of this element to function and the functional difference between V469L and V471L in spite of their spatial proximity.

In summary, MD simulations provided detailed insight into the molecular mechanism underlying the altered function of both variants. Our MD results agree well with the experimentally observed loss-of-function and gain-of-function phenotypes for V469L and V471L, respectively.

## DISCUSSION

Deriving actionable information for patient diagnosis and treatment from clinical sequencing data is a fundamental challenge in genetics and medicine. Current methods for analyzing sequencing data often fail due to the inability to predict the effects of the detected VUS on protein function. Here, we illustrate the power of a “personalized structural biology” pipeline that places candidate VUS into 3D structural models tailored to the patient. The integration of cycles of computational and experimental analysis enabled us to provide mechanistic molecular insights into the different mechanisms by which two proximal candidate *KCNC2* VUSs lead to DEE-like phenotypes. DEE has been previously linked to dysfunction of other ion channels ^58,59^. Moreover, Kv3 channel family members have been previously associated with neurological disorders such as ataxias, epilepsies, schizophrenia, and Alzheimer’s disease ^60^.

The V469L variant occupies the central hydrophobic residue of the PVP motif and the flexibility of this hinge region is critical for channel opening and closing kinetics ^61^. Previous studies in other channels supported this hypothesis, as altering the central hydrophobic residues in Kv1.1 channels from valine to isoleucine, a constitutional isomer of leucine, affected channel kinetics, stability, and conformational dynamics ^50^. The V469L variant resulted in > 50% decrease in peak current and a very slow deactivation with a slight negative shift (∼10mV). Interestingly, the V469L variant caused the Kv3.2 channel to be expressed at < 50% of WT. Molecular dynamics simulations showed that the V469L variant had a smaller pore radius and a higher energetic barrier to ion transfer. The constriction was likely the result of hydrophobic interactions between bulkier L469 residues causing part of the channel pore to become devoid of water molecules ^62,63^.

The V471 variant is immediately adjacent to the PVP hinge motif and resulted in a > 50% increase in the peak current, with moderately slow deactivation, but a drastic negative shift (∼28mV). There was also an increase in protein expression to > 150% of WT and the pore remained fully open in MD and the pore radius for the V471L variant of Kv3.2 channel was slightly wider. An increased number of inter-subunit contacts made by V471L suggests a possible mechanism how this mutant could selectively stabilize the open channel state. The V469L and V471L variants had opposite loss-of-function and gain-of-function effects respectively.

The lower current for V469L versus higher for V471L compared to WT could be explained by the differences in protein expression levels. However, alterations in the protein level would not cause changes in the kinetics of deactivation. The two variants could affect the energy required for the protein to undergo conformational changes between the open and closed states. The adjacency of the variants to the PVP motif would lead to changes to the ability of the helix to kink at the hinge domain and facilitate channel gating. The V469L variant, which results in the central hydrophobic valine of the PVP motif to be substituted by a bulkier leucine extending into the pore, results in a lower tendency for the helix to kink, and therefore, leads to slower channel gating and slower deactivation. In contrast, the stabilization of the V471L channel in the “open” conformation, which would also affect channel gating, is consistent with its moderately slow deactivation. Thus, our results indicate that the variants influence both expression and channel function. However, our results do not identify the cause of the differences in the expression levels of the two variants. It is likely that these result from differences in protein folding or stability. Further studies, such as analysis of protein trafficking in cells, are needed to identify the causes. For experimental simplicity, we carried out our *in vitro* and *in silico* analyses with all four chains carrying the variant of interest. In the future, it would be valuable to evaluate the spectrum of effects for channels carrying different combinations of variant and WT chains; though, we anticipate similar effects.

We have not simulated the entire dynamics involved in channel activation and deactivation. This process is too long to be studied by conventional MD methods. Enhanced MD protocols, which aim at representing the free energy landscape of the molecular system by a set of low-dimensional collective variables, have been used to simulate conformational changes for some ion channel systems ^64^. These methods could be helpful for explaining the observed changes in activation potential and deactivation time. However, no general protocol for deriving a set of collective variables that capture the whole activation and deactivation cycle of Kv channels like Kv3.2 is available yet.

The ability of the channel blocker 4-AP to inhibit the V469L Kv3.2 channel aligned with the loss-of-function phenotype because 4-AP approaches the channel lumen from the cytoplasm ^53,56,57^; therefore the steric hinderance of the pore cavity by V469L should not affect its mechanism of action. Furthermore, the decreased ability of 4-AP to block the V471L channel supports the gain-of-function hypothesis. Channel closing is destabilized in V471L, so 4-AP may not bind as efficiently to the open channel. These results also illustrate how the personalized structural biology approach can help evaluate the effects of possible pharmacological interventions. For example, 4-AP is not likely to help individuals with the V469L variant, it is possible that it could counteract some effects of the V471L variant.

Taken together, the clinical features of our UDN and the reported and the combined experimental and molecular modeling of their *de novo* KCNC2 variants prove their functional role as a cause of DEE. The phenotypes associated with the two variants in *KCNC2* are the result of two fundamentally different molecular mechanisms, even though the residues are only two amino acids away. Analyses of these variants in their structural context was key to revealing their mechanistic and functional heterogeneity. In addition to the contribution to diagnosis, our results also suggest that drugs that modulate the activity of Kv3.2 could be potential treatments. This case study demonstrates the strength of personalized structural biology as a diagnostic method to predict precise molecular hypotheses by taking the context of the variant if interest in the 3D structure of the protein into account. Going forward, this approach has broad applicability across VUS observed in studies from rare disease to cancer.

## METHODS

### Structural modeling of Kv3.2

The tetrameric structural model of human Kv3.2 (UniProtKB accession number: Q96PR1-1, modeled residues: 1-484) was generated by homology modeling using the molecular modeling software suite Rosetta (version 3.10) ^65^. The shaker family voltage dependent potassium channel (Kv1.2-Kv2.1 paddle chimera channel) resolved to 2.4 Å (PDB ID: 2R9R) was used as a template. The percent identity between the aligned positions of the sequences of Kv3.2 and the template structure was 42.8%, sufficiently high for the chimera channel structure to serve as a reliable template. A starting partial tetrameric model of Kv3.2, which only covered aligned residues, was generated by threading the sequence of Kv3.2 onto the template structure using the corresponding sequence alignment as a guide. Full models were created using the Rosetta comparative modeling (RosettaCM) protocol ^66^ guided by the RosettaMembrane energy function ^67^ in a C4 symmetry mode ^68^. The boundaries of membrane-spanning segments were calculated using the PPM server ^69^ based on the starting model. The boundaries were used to impose membrane-specific Rosetta energy terms on residues within the theoretical membrane bilayer. A total of 1000 full tetrameric models were generated using RosettaCM. The lowest-energy model was selected as the final model for structure-based analysis in this work.

### MD system setup

The Kv3.2 channel domain (residues L211 – M484) was embedded in a POPC (palmitoyloleoyl-phosphatidylcholine) bilayer (∼240 lipid molecules per leaflet) using the membrane builder tool of CHARMM-GUI ^70^. The system was solvated in TIP3P water containing 0.15 M of neutralizing KCl. Three K^+^ ions were placed in the channel selectivity filter at coordination sites S0, S2, and S4 by inferring their positions from the crystal structure of the Kv1.2-2.1 chimeric channel (PDB: 2R9R) ^43^. Another K^+^ ion was placed below the selectivity filter in the aqueous channel cavity (termed SCav site) and used for running umbrella simulations. During conventional MD simulations, the position of the cavity K^+^ ion was constrained by imposing distance restraints to the selectivity filter residue T437.

### Conventional MD simulations

MD simulations of the Kv3.2 channel in POPC membranes were performed with AMBER16 ^71^ using the ff14SB force field for proteins ^72^ and the Lipid17 force field. The system was simulated in four replicas with a total simulation time of ∼1 μs. Bonds involving hydrogen atoms were constrained with SHAKE ^73^. Nonbonded interactions were evaluated with a 10 Å cutoff, and long-range electrostatic interactions were evaluated by the particle-mesh Ewald method ^74^. Each MD system was first minimized using a four-step energy minimization procedure: Minimization of only lipids was followed by minimization of only water + ions, and minimization of protein before the whole system was minimized. With protein and ions restrained to their initial coordinates, the lipid and water were heated to 50 K over 1000 steps with a step size of 1 fs in the NVT ensemble using Langevin dynamics with a rapid collision frequency of 10,000 ps^-1^. The system was then heated to 100 K over 50,000 steps with a collision frequency of 1000 ps^-1^ and finally to 310 K over 200,000 steps and a collision frequency of 100 ps^-1^. After changing to the NPT ensemble, restraints on protein and ions were gradually removed over 500 ps. The system was equilibrated for another 10 ns at 310 K with weak positional restraints (with a force constant of 5 kcal mol^-1^ Å^-2^) applied to protein Cα atoms. The protein restraints were then gradually removed over 20 ns, and production MD was conducted for 260 ns using a step size of 2 fs, constant pressure periodic boundary conditions, anisotropic pressure scaling, and Langevin dynamics.

Subsequent to running production MD, molecules were reimaged back into the simulation box using CPPTRAJ ^75^ and the final 200 ns of each MD replica were analyzed. The Kv3.2 channel was aligned to the first MD frame and the channel pore radius was measured with HOLE ^76^ by taking conformations of Kv3.2 at every 1 ns.

### Umbrella MD simulations

In order to estimate the free energy of K^+^ ion permeation through the cytosolic gate of Kv3.2 WT, V469L, and V471L channels, we calculated the potential of mean force (PMF) of moving a K^+^ ion up the pore axis past the cytosolic constriction site and into the cavity below the selectivity filter. The center of mass of the backbone atoms of the selectivity filter residues (T437 – Y440 of all four subunits) was defined as the origin of the pore axis. Umbrella potentials (with a spring constant of 10 kcal mol^-1^ Å^-2^) were placed at 0.5 Å intervals in the range from Z = -11 Å (below selectivity filter) to Z = -44 Å (in cytosol), making a total of 67 umbrella simulations for each 1D PMF. In addition, to ensure that the K^+^ ion remained in the vicinity of the pore axis when it was no longer constrained by the S6 helices (i.e., was in bulk solvent), we used a method described in Fowler et al. ^77^ and applied a flat-bottomed cylindrical constraint with a radius of 8 Å and a spring constant of 10 kcal mol^-1^ Å^-2^. Starting configurations for each umbrella window were prepared by taking the last frame from the conventional MD simulation and gradually pulling the cavity K^+^ ion from SCav into cytosolic solvent over 10 ns using a spring constant of 10 kcal mol^-1^ Å^-2^. To ensure that the direction of the pore axis was well-defined and did not change during the simulation, the positions of the backbone atoms of the first two helix turns of S6 (W448 – G454) were constrained by a harmonic potential with a force constant of 5 kcal mol^-1^ Å^-2^ during the pulling and umbrella simulations. Each umbrella simulation was run for 10 ns and repeated twice for each of the original four MD replicas. The WHAM method ^78^ as implemented in the program by Grossfield ^79^ was used to remove the umbrella biases and calculate 1D PMFs. A final 1D PMF was calculated for Kv3.2 WT, V469L, and V471L, respectively, by averaging the individual PMFs for each variant, and the height of the free energy barrier relative to the bulk solvent was measured.

### Heterologous expression of Kv3.2 ion channel and whole-cell voltage clamp electrophysiology

Wild-type Kv3.2, V469L, and V471L channel plasmids (1 ng/µl for each plasmid) were separately transfected with fluoresced green protein (GFP as marker to identify successful ion channel expression) into Chinese Hamster Ovary (CHO) cells using 10 µl Fugene 6 (Promega), following manufacturer’s cell transfection instructions. Two days post transfection, cells with green color were selected for patch clamp experiments.

Whole-cell voltage clamp experiments were performed at room temperature (22-23°C) with 3∼5 mΩ patch microelectrodes, by using a MultiClamp 700B amplifier and DigiData 1550B low-noise data acquisition system (Molecular Devices Inc., Sunnydale, California). The extracellular solution contained (in mmol/L) NaCl 145, KCl 4.0, MgCl_2_ 1.0, CaCl_2_ 1.8, glucose 10, and HEPES 10; the pH was 7.4, adjusted with NaOH. The pipette (intracellular) solution contained (in mmol/L) KCl 110, MgCl_2_ 1.0, ATP-K_2_ 5.0, BAPTA-K4 5.0, and HEPES 10; the pH was 7.2, adjusted with KOH. Data acquisition was performed using pClamp 10.7 software (Molecular Devices Inc.), sampling at 1 kHz and low-pass-filtered at 5 kHz. Activating current was elicited with 1-second depolarizing pulses from a holding potential of −80 mV at a 10-mV increments, and tail current was recorded on return to −40 mV. The voltage-clamp protocol is shown in Figure EP. Pulses were delivered every 15 seconds. The current-voltage (I-V) relationships were analyzed by fitting the Boltzmann equation to the data: I=I_max_ / {1 + exp [(V_t_ − V_0.5_) / k]}, where I_max_ is the maximal current, V_t_ is the test potential, V_0.5_ is the membrane potential at which 50% of the channels are activated, and k is the slope factor. Current densities (pA/pF) were obtained after normalization to cell surface area calculated by the Membrane Test in pClamp 10.7. A potassium channel blocker 4-aminopyridine (4-AP at 200 µM; Sigma-Aldrich Co., St. Louis, MO, USA) was used to test the sensitivity of wildtype Kv3.2 and two variant (V469L and V471L) channels to drug block by 1-second repetitive pulsing protocol from a holding potential of -80 mV to a testing potential of +60 mV (Figure S3).

### DNA constructs for WT and variants of KCNC2

The coding sequences DNA of human *Homo sapiens* potassium voltage-gated channel subfamily C member 2 *KCNC2* (NM_139137.4)) was subcloned into pcDNA3.1+ /C-(k)-DYK expression vector with an equipped Flag tag (DYKDDDDK) in C-terminal (GenScript, NJ, USA). The mutant KCNC2 variants Kv3.2-V469L and Kv3.2-V471L cDNA constructs were generated by using a pair of designed overlapping primers for the PCR in the QuikChange Site-Directed Mutagenesis Kit (Agilent USA Cat. # 200523) and by PCR Overlap Extension method to introduce the mutation site in. Both variants were confirmed by DNA sequencing.

### Western Blot

To detect the Kv3.2 protein expression and to perform protein functional analysis, the wild type and two mutated cDNA plasmids were transfected to CHO stable cells (ATCC, USA) by X-tremeGENE 9 DNA Transfection Reagent (Roche). The transfected cells were collected and lysed in modified Radio-Immunoprecipitation Assay (RIPA) lysis buffer (50 mM Tris pH = 7.4, 150mM NaCl, 1% NP-40, 0.2% Sodium Deoxycholate, 1mM EDTA), and 1% protease inhibitor cocktail (Sigma-Aldrich Co., USA). Collected protein samples were subjected to gel electrophoresis using 4–12% BisTris NuPAGE precast gels (Invitrogen Life Technologies, USA) and transferred to PVDF-FL membranes (MilliporeSigma, USA). Primary antibody against Flag epitope tag located on FLAG fusion proteins (Sigma-Aldrich, polyclonal ANTI-FLAG, rabbit host. F7425) was used to detect the Kv3.2 protein by indirect immunofluorescent staining at a 1:500 dilution. Anti-Na^+^/K^+^ ATPase antibody (Developmental Studies Hybridoma Bank, Antibodies at the University of Iowa for use in research, USA) at a 1:1000 dilution was used as an internal quality control. IRDye conjugated secondary anti-rabbit antibody (LI-COR Biosciences Inc. USA) was used at a 1:10000 dilution. The membranes were scanned using the Odyssey Infrared Imaging System, and the integrated density value of bands was determined using the Odyssey Image Studio software (LI-COR Biosciences Inc. USA).

### Clinical Report

Please contact the corresponding authors for clinical details.

## Data Availability

All data produced in the present work are contained in the manuscript.

## ACKNOWLEDGEMENTS

First, we thank all affected UDN individuals and their families. We thank members of the Capra, Meiler, and UDN Labs for helpful comments. This work was conducted in part with the resources of the Advanced Computing Center for Research and Education at Vanderbilt University, Nashville, TN.

## AUTHOR CONTRIBUTIONS

Conceptualization, S.M., T.A.C., G.K., J.A.P.III, J.A.C.; Methodology, S.M., B.L., N.H., T.Y., W.S., R.L.M., D.M.R., J.M., G.K., J.A.P.III, J.A.C.; Investigation, S.M., B.L., N.H., T.Y., W.S., J.H.S., C.W.M., G.K.; Writing – Original Draft, S.M., T.A.C., B.L., G.K., N.H., T.Y.; Writing – Review & Editing, S.M., B.L., G.K., J.M., J.A.P.III, G.K., J.A.C; Funding Acquisition, R.H., R.L.M., D.M.R., UDN, J.A.P.III, J.M., J.A.C.; Resources, J.D.C., R.H., J.H.N., R.L.M., D.M.R., J.M., J.A.P.III, J.A.C.; Supervision, D.C.R., R.L.M., D.M.R., J.M., G.K., J.A.P.III, J.A.C.

## CONSORTIUM

### Undiagnosed Diseases Network

Maria T Acosta, David R Adams, Pankaj Agrawal, Mercedes E Alejandro, Patrick Allard, Justin Alvey, Ashley Andrews, Euan A Ashley, Mahshid S Azamian, Carlos A Bacino, Guney Bademci, Eva Baker, Ashok Balasubramanyam, Dustin Baldridge, Jim Bale, Deborah Barbouth, Gabriel F Batzli, Pinar Bayrak-Toydemir, Alan H Beggs, Gill Bejerano, Hugo J Bellen, Jonathan A Bernstein, Gerard T Berry, Anna Bican, David P Bick, Camille L Birch, Stephanie Bivona, John Bohnsack, Carsten Bonnenmann, Devon Bonner, Braden E Boone, Bret L Bostwick, Lorenzo Botto, Lauren C Briere, Elly Brokamp, Donna M Brown, Matthew Brush, Elizabeth A Burke, Lindsay C Burrage, Manish J Butte, John Carey, Olveen Carrasquillo, Ta Chen Peter Chang, Hsiao-Tuan Chao, Gary D Clark, Terra R Coakley, Laurel A Cobban, F Sessions Cole, Heather A Colley, Cynthia M Cooper, Heidi Cope, William J Craigen, Precilla D’Souza, Surendra Dasari, Mariska Davids, Jyoti G Dayal, Esteban C Dell’Angelica, Shweta U Dhar, Naghmeh Dorrani, Daniel C Dorset, Emilie D Douine, David D Draper, Laura Duncan, David J Eckstein, Lisa T Emrick, Christine M Eng, Cecilia Esteves, Tyra Estwick, Liliana Fernandez, Carlos Ferreira, Elizabeth L Fieg, Paul G Fisher, Brent L Fogel, Irman Forghani, Laure Fresard, William A Gahl, Rena A Godfrey, Alica M Goldman, David B Goldstein, Jean-Philippe F Gourdine, Alana Grajewski, Catherine A Groden, Andrea L Gropman, Melissa Haendel, Neil A Hanchard, Nichole Hayes, Frances High, Ingrid A Holm, Jason Hom, Yong Huang, Alden Huang, Rosario Isasi, Fariha Jamal, Yong-Hui Jiang, Jean M Johnston, Angela L Jones, Lefkothea Karaviti, Emily G Kelley, Dana Kiley, David M Koeller, Isaac S Kohane, Jennefer N Kohler, Susan Korrick, Mary E Koziura, Deborah Krakow, Donna M Krasnewich, Joel B Krier, Jennifer E Kyle, Seema R Lalani, Byron Lam, Brendan C Lanpher, Ian R Lanza, C Christopher Lau, Pace Laura, Jozef Lazar, Kimberly LeBlanc, Brendan H Lee, Hane Lee, Roy Levitt, Shawn E Levy, Richard A Lewis, Sharyn A Lincoln, Pengfei Liu, Xue Zhong Liu, Nicola Longo, Sandra K Loo, Joseph Loscalzo, Richard L Maas, Calum A MacRae, Ellen F Macnamara, Valerie V Maduro, Marta M Majcherska, May Christine V Malicdan, Laura A Mamounas, Teri A Manolio, Rong Mao, Thomas C Markello, Ronit Marom, Gabor Marth, Beth A Martin, Martin G Martin, Julian A Martínez-Agosto, Shruti Marwaha, Thomas May, Jacob McCauley, Allyn McConkie-Rosell, Colleen E McCormack, Alexa T McCray, Thomas O Metz, Matthew Might, Eva Morava-Kozicz, Paolo M Moretti, Marie Morimoto, John J Mulvihill, David R Murdock, Avi Nath, Stanley F Nelson, J Scott Newberry, Sarah K Nicholas, Donna Novacic, Devin Oglesbee, James P Orengo, Stephen Pak, J Carl Pallais, Christina G S Palmer, Moretti Paolo, Jeanette C Papp, Neil H Parker, Jennifer E Posey, John H Postlethwait, Lorraine Potocki, Barbara N Pusey, Aaron Quinlan, Archana N Raja, Genecee Renteria, Chloe M Reuter, Lynette C Rives, Amy K Robertson, Lance H Rodan, Jill A Rosenfeld, Robb K Rowley, Maura Ruzhnikov, Ralph Sacco, Jacinda B Sampson, Susan L Samson, Mario Saporta, Judy Schaechter, Timothy Schedl, Kelly Schoch, Daryl A Scott, Lisa Shakachite, Prashant Sharma, Vandana Shashi, Kathleen Shields, Jimann Shin, Rebecca H Signer, Catherine H Sillari, Edwin K Silverman, Janet S Sinsheimer, Kathy Sisco, Kevin S Smith, Lilianna Solnica-Krezel, Rebecca C Spillmann, Joan M Stoler, Nicholas Stong, Jennifer A Sullivan, Shirley Sutton, David A Sweetser, Holly K Tabor, Cecelia P Tamburro, Queenie K-G Tan, Mustafa Tekin, Fred Telischi, Willa Thorson, Cynthia J Tifft, Camilo Toro, Alyssa A Tran, Tiina K Urv, Matt Velinder, Dave Viskochil, Tiphanie P Vogel, Colleen E Wahl, Melissa Walker, Nicole M Walley, Chris A Walsh, Jennifer Wambach, Jijun Wan, Lee-Kai Wang, Michael F Wangler, Patricia A Ward, Katrina M Waters, Bobbie-Jo M Webb-Robertson, Daniel Wegner, Monte Westerfield, Matthew T Wheeler, Anastasia L Wise, Lynne A Wolfe, Jeremy D Woods, Elizabeth A Worthey, Shinya Yamamoto, John Yang, Amanda J Yoon, Guoyun Yu, Diane B Zastrow, Chunli Zhao, Stephan Zuchner.

## SUPPLEMENTARY FIGURES

**FIGURE S1:**
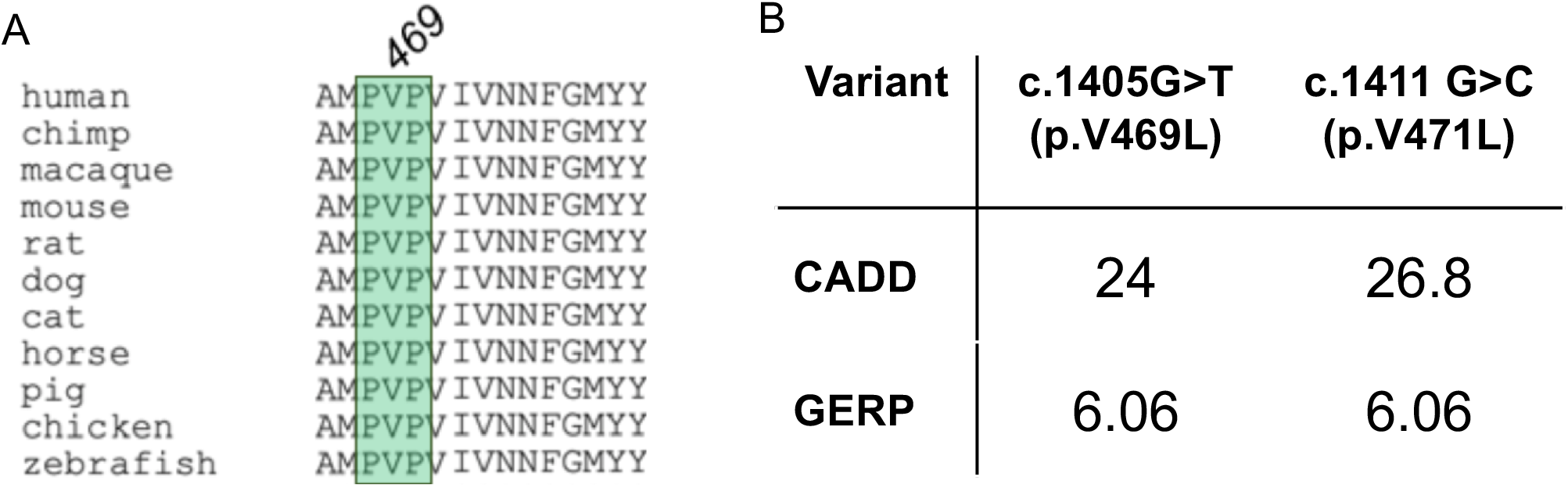
*KCNC2* variants are located in the evolutionarily conserved hinge domain and predicted to be deleterious. **(A)** The amino acid sequence flanking the PVP motif (green box) and the V469L and V471L are deeply conserved across vertebrate species. **(B)** V469L and V471L are both predicted to be functional by the combined annotation dependent depletion (CADD) and genomic evolutionary rating profile (GERP) scores.

**FIGURE S2:**
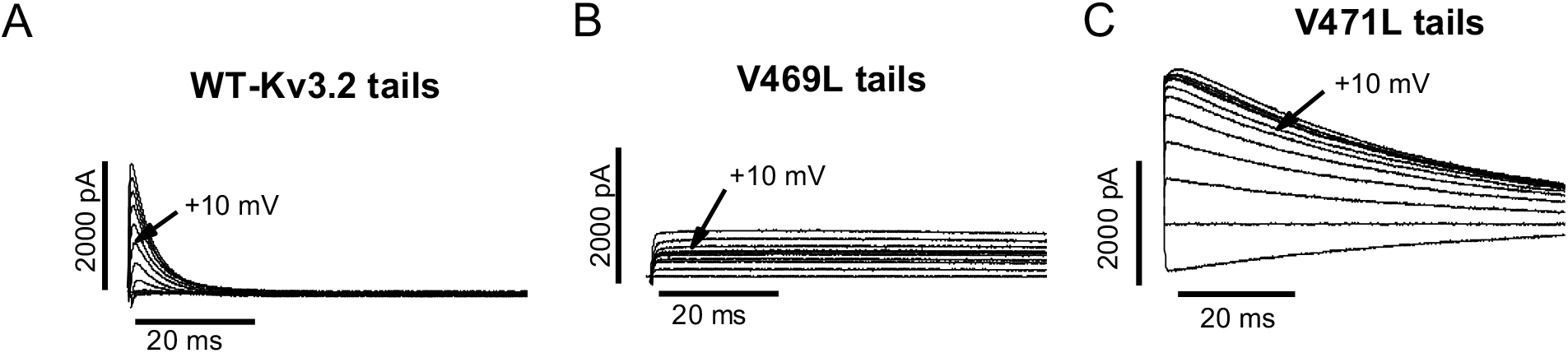
Candidate *KCNC2* variants have different effects on Kv3.2 deactivation tail kinetics. **(A)** Current traces for the deactivating tails for wild type (WT) Kv3.2. The short peak indicates fast deactivation, which is a hallmark of Kv3.2, and required for fast depolarization of membrane potential in the central nervous system. **(B)** Current traces for the deactivating tails of V469L Kv3.2 show a much slower deactivation, indicated by the long peak which does not return to zero. **(C)** Current traces for the deactivating tails of V471L Kv3.2 demonstrate a much higher current and a somewhat slowed deactivation, indicated by the higher and longer peak, compared to the WT.

**FIGURE S3:**
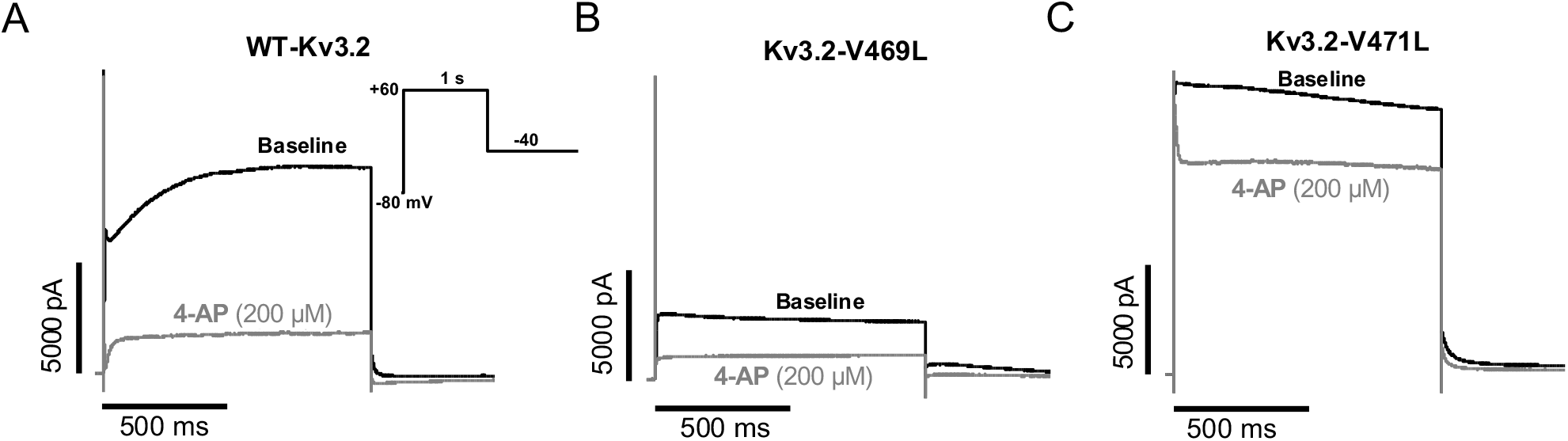
4-AP differentially blocks variant Kv3.2 channels. The effect of 4-aminopyridine (4-AP), a voltage gated potassium channel blocker, on WT Kv3.2 **(A)**, V469L **(B)**, and V471L **(C)** channel activity. Representative baseline (absence of 4-AP) current traces are shown in black for each protein, and representative traces with 200 uM 4-AP are shown in gray. 4-AP blocked the WT and V469L at similar levels. However, activity of the V471L variant was only modestly reduced by 4-AP and maintained high activity. The voltage-clamp protocol is shown as insert in panel A.

**FIGURE S4:**
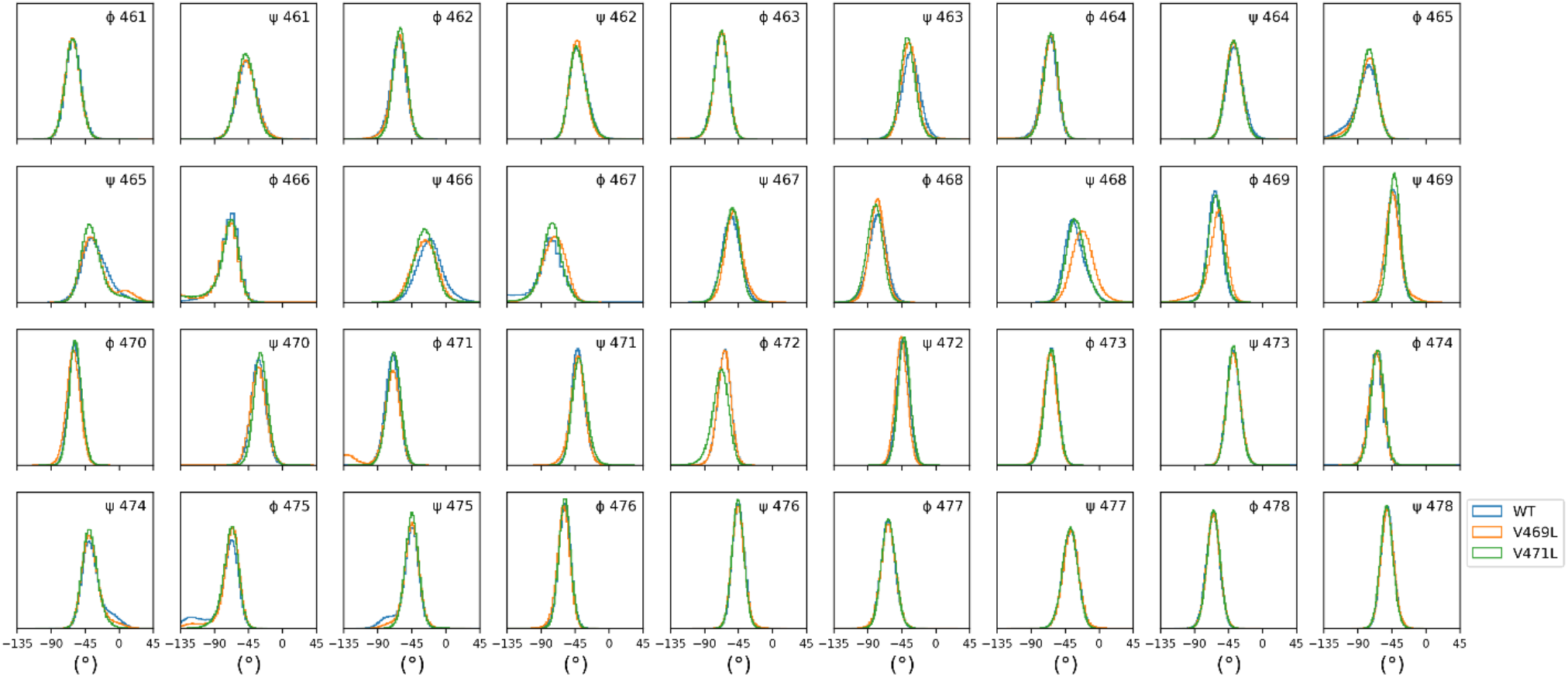
Distribution of ϕ and Ψ backbone angles of pore helix residues 461 to 478 sampled in MD simulation of Kv3.2 WT, V469L, and V471L. For Kv3.2 V469L variant, small changes are found at Ψ 466, ϕ 467, Ψ 468 and ϕ 469, and for Kv3.2 V471L at Ψ 466 (**TABLE T1**)

**TABLE T1:**
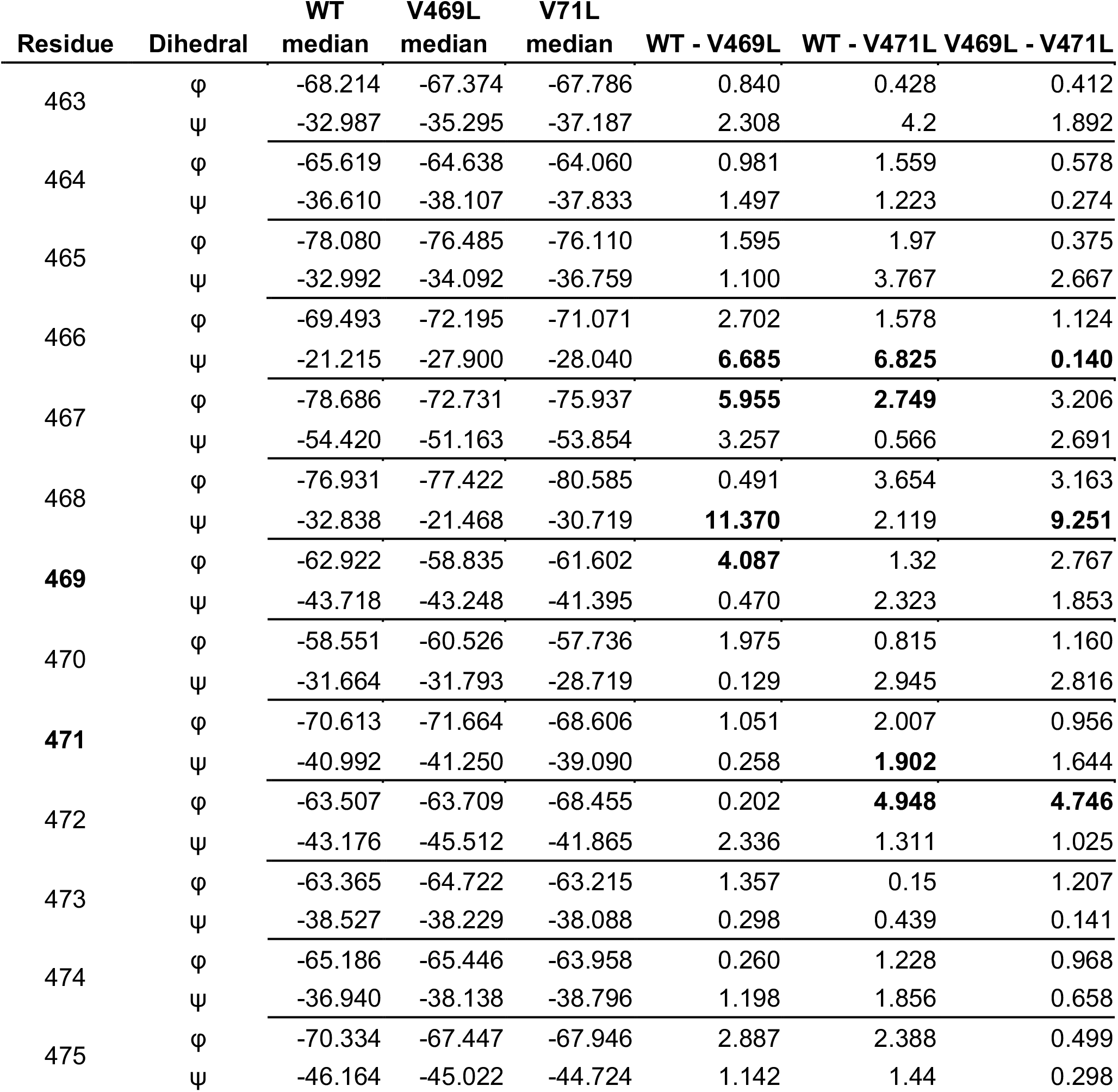
Summary of ϕ and Ψ backbone angles of pore helix residues 463 to 475 sampled in MD simulation of Kv3.2 WT, V469L, and V471L. For Kv3.2 V469L variant, small changes are found at Ψ and ϕ angles for several residues. The alterations for Ψ 466 were consistent between both the variants, indicating a strain on the S6 helix resulting from both variants. However, the difference between V469L and WT for Ψ 468 and ϕ 469 were specific for V469L variant, and not observed for the V471L variant. Conversely, the alterations to Ψ 471 and ϕ 472 were observed specifically for the V471L variant, indicating these changes were specific for the position and resulting amino acid of the variants.

**FIGURE S5:**
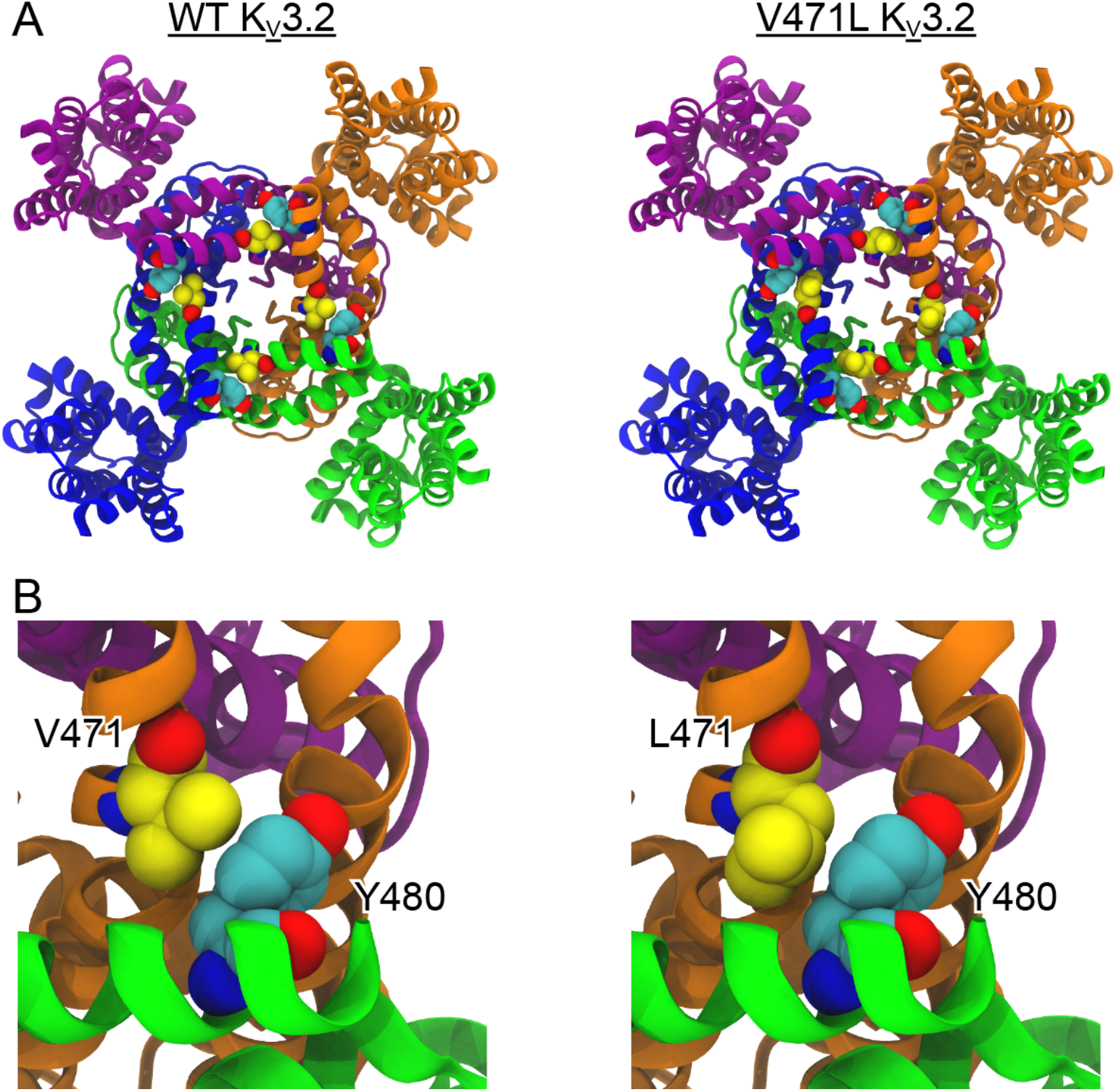
Adjacent pore helices in Kv3.2 form a contact between residues 471 and 480. **(A)** Cytosolic view of structural models of Kv3.2 WT and V471L. Each subunit is shown with a different color. V471 in WT Kv3.2 and L471 in the pV471L variant, respectively, are represented as spheres and colored yellow. Residue Y480 is shown in cyan. **(B)** Close-up view of residues V471 (left) or L471 (right) interacting with Y480.

